# Tirzepatide for Lipodystrophy

**DOI:** 10.1101/2024.09.25.24313345

**Authors:** Rasimcan Meral, Merve Celik Guler, Diarratou Kaba, Jeevitha Prativadi, Eric D Frontera, Maria Cristina Foss-Freitas, Noura Nachawi, David T Broome, Marissa Lightbourne, Rebecca J Brown, Simeon I Taylor, Elif A Oral

## Abstract

**Background:** Lipodystrophy encompasses a group of rare disorders associated with severe metabolic disease. These disorders are defined by abnormal fat distribution, with near-total (generalized lipodystrophy, GL) or partial (partial lipodystrophy, PL; i.e. familial partial lipodystrophy, FPLD) absence of adipocyte mass leading to a decreased ability to store lipids safely. Excess lipids are more likely to be stored in non-adipose tissues, which leads to the metabolic manifestations. We have recently shown that glucagon-like peptide-1 agonists are associated with metabolic improvements in FPLD. We hypothesized that tirzepatide, a dual incretin, may also lead to metabolic improvement in patients with lipodystrophy.

**Methods:** Observational cohort of patients with PL or GL who received tirzepatide clinically were tracked in the context of ongoing natural history studies.

**Results:** Seventeen patients received tirzepatide, 14 with FPLD (ages within 30-74 years; 12 female 2 male). After a median 8.7 months of follow-up, BMI (medianΔ -1.7; range -5.9 to 0.9 kg/m^2^; *p*=0.008), HbA1c (medianΔ -1.1%; range -6.3 to -0.1%; *p*<0.001), triglycerides [medianΔ - 65 mg/dL (-0.73 mmol/L); range -3820 to 43 mg/dL (-43.2 to 0.49 mmol/L); *p*=0.003] and total daily insulin requirements (medianΔ -109; range -315 to 0 units/day; *p*=0.002) were significantly reduced. Three patients with acquired GL (Ages within 35-64 years; all female) also demonstrated a robust response to tirzepatide with reduced BMI (22.2->20.9; 26.2->25.4; 19.5->17.6 kg/m^2^), HbA1c (8.5%->7.0%; 10.2%->7.8%; 9.1%->6.5%), triglycerides (91->80; 641->293; 1238->100 mg/dL or 1.03->0.90; 7.24->3.31; 14.0->1.13 mmol/L), and total daily insulin requirement (85->0; 0->0; 1000->750 units/day). Three patients did not tolerate dose escalation due to gastroesophageal reflux.

**Conclusions:** Tirzepatide may be an effective treatment for patients with lipodystrophy.

## Introduction

Lipodystrophy encompasses a group of disorders defined by abnormal distribution of fat. Generalized lipodystrophy (GL) is characterized by near-total absence of adipocyte mass and function whereas partial lipodystrophy (PL) is characterized by a partial decrease in adipocyte mass and function.^1,2^ Patients describe profound hyperphagia secondary to hypoleptinemia.^3,4^ Hyperphagia coupled with reduced ability to store excess calories safely within adipocytes leads to ectopic spill-over of lipids, leading to the severe metabolic sequelae observed.^5^ These include, but are not limited to, severe insulin resistance, diabetes mellitus and its complications, severe hypertriglyceridemia with recurrent pancreatitis, and metabolic dysfunction associated steatohepatitis (MASH) that can progress to end-stage liver disease.^6-10^ Therapeutic agents with potent anorexigenic effects are of great interest in this population.

Metreleptin is an injectable peptide initially studied in the 1990’s as a drug for obesity but failed in trials due to lack of substantial efficacy in hyperleptinemic obesity.^11,12^ It was, however, remarkably effective in patients with severe forms of lipodystrophy and severely reduced leptin levels^13-17^ culminating in its approval for the treatment of GL by the FDA in 2014^18^ and by the EMA in 2018.^19^ In contrast, response to metreleptin has been variable in patients with PL who have selective absence of major fat depots and are not always leptin deficient.^20-22^ PL remains a condition with severe metabolic disease burden, intractable to conventional therapeutics and with no approved treatment options in the US. While a global randomized controlled study with a different therapeutic targeting APO-CIII (volenasorsen) led to significant sustained reduction in hypertriglyceridemia and hepatic statosis in PL, side effects of thrombocytopenia and increased systemic inflammation limit the broad applicability with approval granted only in Brazil.^23^

Incretin mimetics have revolutionized clinical obesity management through central and peripheral effects to ameliorate glucose metabolism.^24^ We became interested in the potential impact of incretins because the main benefit of leptin replacement are thought to be driven by reduced caloric intake through induced satiety^3,13,25^ and alternative agents to lower caloric intake may thus be impactful. Furthermore, we noted rapid and remarkable metabolic improvement in a single case of familial partial lipodystrophy (FPLD) with very-low-calorie diet, highlighting the importance of controlling food intake in this population.^26^ We recently described the clinical efficacy of pure GLP-1 agonists in a retrospective cohort of 14 patients with PL, which remains the largest case evidence that these drugs may exert beneficial metabolic effects besides a few case reports.^27-29^ Tirzepatide is a dual glucose-dependent insulinotropic polypeptide/glucagon-like-peptide-1 receptor agonist (GIP/GLP1-RA) that induces dramatic weight loss in individuals with obesity while also improving diabetes and other metabolic complications.^30-32^ It is currently the most efficacious approved drug for managing obesity. We hypothesized that tirzepatide would ameliorate metabolic disease burden in patients with lipodystrophy. To that end, we collected data from patients who are being prescribed these medications who are also part of our ongoing prospective natural history studies. This report summarizes data captured from these efforts.

### Research Design and Methods

This is an observational cohort study involving patients with lipodystrophy followed by either University of Michigan (n=14) or National Institute of Diabetes and Digestive and Kidney Diseases (NIDDK) Intramural Program (n=3) who were prescribed tirzepatide for the management of their diabetes and/or weight. Data were gathered as a part of ongoing natural history studies at each site, with details of each study in **Table S1**. Patients who received at least one dose of tirzepatide between November 2022 and March 2024 were included. Patients who were still on treatment but lacked follow-up data were excluded, but those who failed treatment for any reason were included. BMI, Hemoglobin A1c, triglycerides and total daily insulin requirement before and after tirzepatide were compared. Follow-up visit and lab frequency, dose escalations, symptom management and concurrent medication changes while on tirzepatide were at physicians’ discretion and occurred independent from inclusion in this study. Patients were advised on the potential gastrointestinal side effects to be expected with tirzepatide use, but did not receive any dietary advice aimed at caloric restriction. Most recent vitals and labs prior to the first dose of tirzepatide were used as baseline data. As follow-up data, most recent vitals and labs while the patient was still on the drug were used.

Only patients with PL are included in descriptive and inferential statistics; patients with GL are discussed as individual cases. Before and after data were compared using Wilcoxon matched-pairs signed rank test. Data are reported *median (minimum-maximum)* or *median [inter-quartile-range]* . Ages of individual patients are reported as ranges containing the patients’ age for deidentification purposes. A Bonferroni correction for four comparisons was made and P < 0.0125 was considered statistically significant. All tests were performed using GraphPad Prism v10.2.2 (Boston, Massachusetts).

## Results

A total of 17 patients with lipodystrophy (14 with PL, 3 with GL) were treated with tirzepatide. Fifteen were prescribed tirzepatide to improve glycemic control, patient #05 and #11 were prescribed tirzepatide for weight management. As with lipodystrophy overall, this study included a very heterogeneous population, reflected in a range of comorbidities (**Table S2**) and a wide array of background medications (**Table S3**). Confounding medication changes during the observed period were rare. Most patients (9 of 17) had previously tried and discontinued other GLP-1 RA or dipeptidyl-peptidade-4 inhibitors (**Table 1**). Three of these transitioned directly from another GLP-1 RA to tirzepatide. The reason for change was either room for improvement in glycemic control or tolerance issues as described in **Table S4**. At the time of tirzepatide initiation, 5 patients were treated with metreleptin. Metreleptin was continued unchanged for 3 patients due to lack of efficacy, but discontinued for the other two.

**Table 1:** Baseline characteristics of patients. *Age is reported as age range containing the patients’ age for deidentification purposes §: Sex assigned at birth was female. Patient identifies as a male and is currently undergoing gender corrective treatment. †: FPLD1 is defined as FPLD without confirmed genetic cause. This patient has a pathogenic variant confirmed to be associated with lipodystrophy, but there is no other subtype yet defining it. ‡: Metreleptin was discontinued tirzepatide was initiated during the same clinic visit. AGL, Acquired Generalized Lipodystrophy; DPP-4, Dipeptidyl Peptidase-4; EA, Expanded Access program; FPLD1, FPLD2, FPLD3 represent Familial Partial Lipodystrophy type 1, 2, and 3, respectively; GERD, Gastroesophageal Reflux Disease; GLP-1RA, Glucagon-Like Peptide-1 Receptor Agonist; JDM, Juvenile Dermatomyositis; NA, Not Applicable; NIH, National Institutes of Health; UM, University of Michigan.

Figures
|  | Site | Age range* (year) | Sex | Race and ethnicity | Subtype | Pathogenic variants | Previous GLP-1RA or DPP-4 inhibitor use | Concurrent leptin replacement |
| --- | --- | --- | --- | --- | --- | --- | --- | --- |
| #01 | UM | 40-44 | female | white, non-hispanic | FPLD1 | *PPP1R3A* c.1985_1986delAG† | - | ‡ (EA) |
| #02 | UM | 50-54 | female | white, non-hispanic | FPLD2 | *LMNA* c.1444C>T (p.R482W) | liraglutide, semaglutide‡ | - |
| #03 | UM | 30-34 | female | white, non-hispanic | FPLD1 | - | liraglutide | - |
| #04 | UM | 40-44 | female | white, non-hispanic | FPLD2 | *LMNA* c.1445G>A (p.R482Q) | - | - |
| #05 | UM | 45-49 | male | white, non-hispanic | FPLD1 | - | liraglutide | - |
| #06 | UM | 35-39 | female | other, hispanic | FPLD3 | *PPARG* c.528_531dup (p.L178*) | dulaglutide‡ | - |
| #07 | UM | 40-44 | female | white, non-hispanic | FPLD2 | *LMNA* c.1445G>T (p.R482L) | exenatide, liraglutide, dulaglutide | ‡ (EA) |
| #08 | UM | 20-24 | female§ | white, non-hispanic | FPLD1 | - | exenatide, dulaglutide, liraglutide, semaglutide | - |
| #09 | UM | 40-44 | female | white, non-hispanic | FPLD1 | - | dulaglutide | - |
| #10 | UM | 55-59 | female | white, non-hispanic | FPLD2 | *LMNA* c.1445G>A (p.R482Q) | dulaglutide, liraglutide, semaglutide | Yes (EA) |
| #11 | UM | 40-44 | female | white, non-hispanic | FPLD2 | *LMNA* c.178C>G (p.R60G) | - | Yes |
| #12 | UM | 70-74 | male | white, non-hispanic | FPLD1 | - | liraglutide, semaglutide‡ | - |
| #13 | NIH | 35-39 | female | white, non-hispanic | FPLD1 | - | - | - |
| #14 | NIH | 40-44 | female | white, non-hispanic | FPLD2 | *LMNA* c.1444C>T (p.R482W) | - | - |
| GL1 | UM | 50-54 | female | black, non-hispanic | JDM-AGL | - | - | - |
| GL2 | UM | 60-64 | female | white, non-hispanic | AGL | - | - | - |
| GL3 | NIH | 35-39 | female | white, non-hispanic | JDM-AGL | - | semaglutide | Yes |

### Response in Partial Lipodystrophy

Fourteen patients with FPLD were included in the study. Demographic information is provided in **Table 1**. Follow-up duration ranged from 3.0-17.4 months (**Table 2**). At the end of the follow-up, reductions in BMI (median difference -1.7 kg/m^2^; range -5.9 to 0.9 kg/m^2^; *p*=0.008), A1c (median difference -1.1%; range -6.3 to -0.1%; *p*<0.001), triglycerides [median difference -65 mg/dL (-0.73 mmol/L); range -3820 to 43 mg/dL (-43.2 to 0.49 mmol/L); *p*=0.003], and total daily insulin dose (median difference -109 units/day; range -315 to 0 units/day; *p*=0.002) were observed (**Figure 1**). Caloric intake was not measured, but all patients verbalized marked reductions in daily food intake.

**Table 2:** Before and after metabolic parameters of all patients. *: Patient underwent left sided below knee amputation 6 months after treatment initiation. AGL, Acquired Generalized Lipodystrophy; FPLD, Familial Partial Lipodystrophy; IQR, Inter-Quartile Range; NA, Not Applicable.

|  | Follow-up<br>(months) | Tolerated<br>tirzepatide<br>(mg/week) | BMI<br>(Kg/m<sup>2</sup>) |  | Hemoglobin A1c<br>(%) |  | Triglycerides<br>[mg/dL (mmol/L)] |  | Insulin use<br>(units/day) |  |
| --- | --- | --- | --- | --- | --- | --- | --- | --- | --- | --- |
|  |  |  | Before | After | Before | After | Before | After | Before | After |
| #01 | 7.0 | 15.0 | 25.0 | 24.5 | 13.4 | 7.1 | 539 (6.1) | 339 (3.8) | 160 | 50 |
| #02 | 17.4 | 12.5 | 30.6 | 28.0 | 7.1 | 6.8 | 164 (1.9) | 156 (1.8) | 82 | 18 |
| #03 | 11.0 | 15.0 | 37.7 | 33.5 | 7.6 | 5.2 | 367 (4.1) | 196 (2.2) | 0 | 0 |
| #04 | 7.9 | 5.0 | 27.6 | 23.4 | 5.4 | 5.1 | 388 (4.4) | 101 (1.1) | 0 | 0 |
| #05 | 13.7 | 15.0 | 37.7 | 38.3 | 8.9 | 8.2 | 459 (5.2) | 394 (4.4) | 280 | 160 |
| #06 | 4.9 | 7.5 | 24.0 | 24.9 | 9.7 | 6.8 | 4080 (46.1) | 259 (2.9) | 124 | 111 |
| #07 | 12.6 | 2.5 | 33.4 | 31.7 | 8.6 | 8.5 | 150 (1.7) | 172 (1.9) | 375 | 60 |
| #08 | 3.0 | 7.5 | 29.9 | 29.2 | 7.9 | 6.8 | 129 (1.5) | 172 (1.9) | 141 | 32 |
| #09 | 11.0 | 2.5 | 46.6 | 47.0* | 7.3 | 6.0 | 309 (3.5) | 261 (2.9) | 378 | 228 |
| #10 | NA | NA | 21.8 | NA | 7.2 | NA | 165 (1.9) | NA | 0 | NA |
| #11 | 5.5 | 2.5 | 18.2 | 16.5 | 6.6 | 6.4 | 298 (3.4) | 117 (1.3) | 90 | 48 |
| #12 | 9.5 | 5.0 | 25.7 | 24.6 | 5.8 | 5.4 | 299 (3.4) | 235 (2.7) | 0 | 0 |
| #13 | 4.8 | 10.0 | 31.6 | 25.7 | 6.8 | 5.7 | 211 (2.4) | 116 (1.3) | 150 | 33 |
| #14 | 11 | 15.0 | 36.1 | 31.9 | 7.6 | 5.8 | 109 (1.2) | 71 (0.8) | 270 | 0 |
| **median [IQR]** | **8.7 [6.0]** |  | **30.3 [10.3]** | **28.0 [7.3]** | **7.5 [1.6]** | **6.4 [1.1]** | **299 [219]**<br>**(3.4 [2.5])** | **172 [142]**<br>**(1.9 [1.6])** | **133 [222]** | **33 [60]** |
| GL1 | 3.5 | 5.0 | 22.2 | 20.9 | 8.5 | 7.0 | 91 (1.0) | 80 (0.9) | 85 | 0 |
| GL2 | 8.9 | 5.0 | 26.2 | 25.4 | 10.2 | 7.8 | 641 (7.2) | 293 (3.3) | 0 | 0 |
| GL3 | 7.4 | 10.0 | 19.5 | 17.6 | 9.1 | 6.5 | 1240 (14.0) | 100 (1.1) | 1000 | 750 |

**Figure 1:**
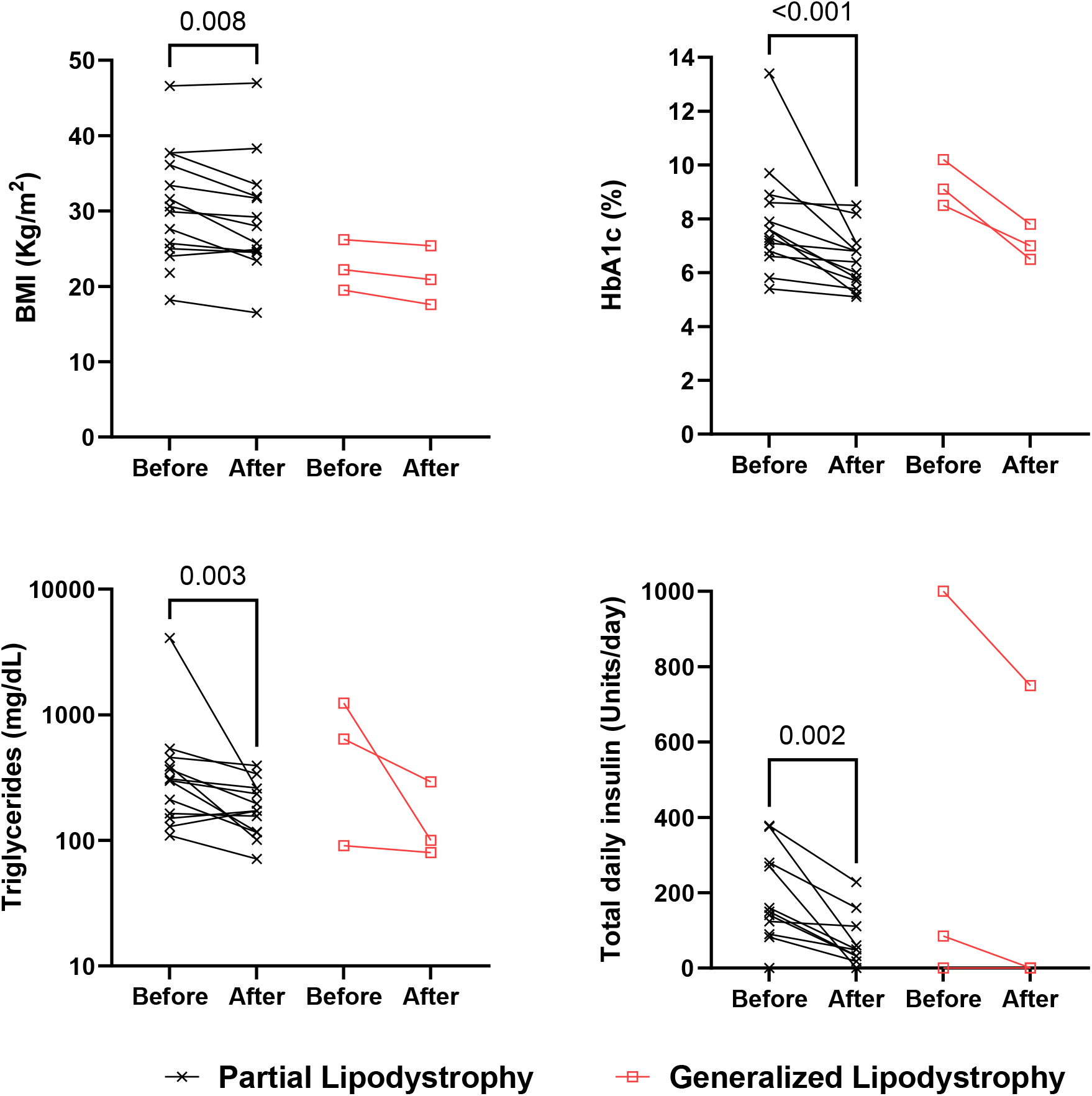
Changes in metabolic parameters before and after tirzepatide. BMI, Body Mass Index; 1c, Hemoglobin A1c.

Ten of 14 patients tolerated tirzepatide at doses shown to be efficacious to treat diabetes and obesity (≥5.0 mg/week). Gastrointestinal side effects including nausea and worsening gastroesophageal reflux disease (GERD) were common and prohibited dose escalation in 3 patients (**Table S4**). Patient #06 experienced episodes of hypertriglyceridemia induced pancreatitis after abrupt cessation of tirzepatide due to national drug shortages (**Figure S2**). Incidentally observed hypoglycemia in patients on concomitant insulin therapy prompted aggressive down-titration of insulin in all patients. One patient (#9) had diabetic foot ulcers at baseline which were complicated by infection leading to a below-knee amputation during this study period, unlikely to be related to tirzepatide.

### Response in Generalized Lipodystrophy

**GL1** is a female with Juvenile Dermatomyositis associated AGL had previously tried metreleptin but without clinical success, which was attributed to challenges in medication adherence. To address her poor glycemic control and to assist with dietary efforts, tirzepatide was chosen as the next treatment; the simpler once weekly dosing was desired. Within 3 months, she lost weight (53.5 to 50.3 kg, BMI 22.2 to 20.9 kg/m^2^), her HbA1c improved (8.5 to 7.0%) and she discontinued insulin (85 to 0 units/day). Her triglycerides were low but still decreased and after tirzepatide (91 to 80 mg/dL). She discontinued tirzepatide after 3 months due to negative cosmetic impact of weight loss on her facial appearance. Six months after discontinuation her BMI increased to 21.9 kg/m^2^, HbA1c increased to 8.6%, and daily insulin requirement rose to 40 units/day.

**GL2** is a female diagnosed with AGL of unknown etiology between 5-9 years of age. She initiated tirzepatide due to inadequate glycemic control with metformin and sulfonylurea. At 6 months follow-up, her BMI decreased from 26.2 Kg/m^2^ to 25.4 Kg/m^2^ and HbA1c improved from 10.2% to 7.8%. At 9 months of follow-up, her triglycerides improved from 641 to 293 mg/dL (7.2 to 3.3 mmol/L). As a potential confounder, she was started on fenofibrate 2 weeks before the first dose of tirzepatide, which may partially explain the triglyceride reduction but is an unlikely explanation for improved in BMI and HbA1c.

**GL3** is a female with Juvenile Dermatomyositis associated AGL who had severe insulin resistance (HbA1c 9.1% despite insulin 1000 units/day and maximal dosing of metreleptin [10 mg/day]). Semaglutide was added for diabetes control but was not tolerated due to nausea. She was therefore switched from semaglutide to tirzepatide. After 7.5 months, her HbA1c decreased to 6.5% and her insulin requirement decreased by 25%. Her triglycerides were reduced by 91% (1240 to 100 mg/dL or 14.0 to 1.1 mmol/L). These changes were associated with weight loss (BMI 19.5 to 17.6 Kg/m^2^).

## Discussion

This study provides observational data in a heterogeneous cohort of 17 patients with PL and GL who were treated with tirzepatide for their clinical care. While this study is not a randomized clinical trial, the breadth of our data in these rare, clinically challenging, and heterogeneous conditions provides the first evidence that this drug may offer substantial clinical benefit in lipodystrophy. In patients with PL, we observed an absolute 1.1% reduction in HbA1c (*p*<0.001) despite a mean insulin reduction of 101 IU/day (*p*=0.003). Tirzepatide was also associated with a 1.9 kg/m^2^ BMI reduction (*p*= 0.008) and clinically meaningful improvement in triglycerides (median reduction of 35% from baseline, *p*=0.003). While our original intent was to review the effects noted in patients with PL, we then expanded the study to include those patients with GL who were also prescribed this medication. The metabolic improvements noted in individuals with GL were also robust and, in our opinion, clinically impactful.

In normal physiology, excess lipids are safely contained within adipocytes. Lipodystrophy syndromes constitute a human model for inadequacy of adipocyte lipid storage, suggesting that the more hazardous aspect of obesity (or increased adiposity) is not high adipocyte mass per se, but rather the inability to further expand fat mass due to having reached the total body adipocyte storage limit, causing a state of adipocyte failure in the face of ongoing excess caloric intake.^5^ With the adipocytes strained, agents such as insulin fail to drive calories into adipocytes, which then results in ectopic tissue deposition. These may thus be considered as mere temporizing efforts, while reducing total body lipids in both obesity and lipodystrophy requires daily caloric deficit. In lipodystrophy, the key to addressing the underlying cause of metabolic complications is to ‘quench’ the hyperphagia caused by the leptin deficient state. This was first achieved with metreleptin, which failed as an obesity drug^11,12^ but proved to be a breakthrough therapy for GL.^13-17,33^ Two decades later, comparable effects may be achieved using incretin mimetic therapies, this time for both GL and PL alike.

As mentioned previously, metreleptin is approved for GL in the US, but not for PL. The approval in EU, UK, Brazil and Japan also covers patients with PL who are not well controlled with other metabolic therapies. In patients with GL, metreleptin can lower HbA1c by 2.0%, triglycerides by 55%^34^, and liver volume by 34%.^17^ In addition, proteinuria was improved,^35^ and female patients achieved a more normal reproductive cycle.^36^ More recently, a survival benefit was also demonstrated.^8^ The improvements noted in patients with PL have remained variable and more modest suggesting that there was not a uniform response (-0.6% reduction in HbA1c, -21% reduction in triglyceride levels).^21^ In our retrospective evaluation of the GLP-1 agonists in patients with PL, we saw absolute 0.5% reduction in HbA1c and a variable response in triglyceride levels while noting 4% reduction in body weight.^29^ These numbers put the improvements noted in this paper into perspective and suggest that tirzepatide has comparatively stronger effects on the PL population.

The small number of patients with GL do not allow for definitive comparisons. However, the large magnitude of the observed changes suggest that the effects are likely real. Metreleptin may not fully address metabolic issues in all patients with GL which suggests that tirzepatide may be a promising therapeutic for patients with GL who do not respond adequately to metreleptin. This expectation is further supported by a recent study of a mouse model of GL using liraglutide, a much weaker pure GLP-1 agonist.^37^ We observed two patients (1 PL and 1 GL) who were on concurrent treatment with tirzepatide and metreleptin. Both demonstrated a robust response to tirzepatide while having no adverse effects. The cotreatment led to a more robust reduction in food intake, weight loss and preservation of lean mass. This is consistent with rodent data from the Myers lab that demonstrates that the central satiety effect of leptin may be mediated by neurons that co-express GLP-1 and leptin receptor.^38^ In their hands, cotreatment led to a more robust reduction in food intake, weight loss and preservation of lean mass. A potential synergy between incretin and leptin pathways warrants further investigation.

Most patients studied in this paper lost weight, and seven of 14 patients with PL lost >5 percent of their body weight. We also noted some weight loss in patients with GL. We are unable to confirm whether the weight was lost from adipose tissue compartments, ectopic lipid storage depots (i.e. liver), or lean mass as body composition was not measured. Studies show that GLP-1 agonists and tirzepatide improve hepatic steatosis.^32,39^

Overall, tirzepatide was tolerated well with adverse reactions that are consistent with the known side effects included in the package insert and mostly considered mild to moderate. One patient experienced a pancreatitis episode when she acutely discontinued tirzepatide due to supply chain shortages. This patient was one of the cases with the highest triglyceride levels in the cohort and achieved a remarkable reduction with tirzepatide. This case may suggest that tirzepatide should be withdrawn with a tapering plan rather than acutely discontinued in patients prone to severe hypertriglyceridemia. One patient with acquired generalized lipodystrophy, who was otherwise doing well, found the cosmetic impact of facial fat loss unacceptable and tirzepatide was stopped.

Lipodystrophy is a BMI independent state, and definitions of normal weight are quite different. GL represents exceptionally low BMI’s and we have previously reported that patinets with PL carry a metabolic burden equivalent to that of controls that were 8-12 kg/m^2^ heavier.^40^ This highlights that BMI should not be used to determine treatment threshold with tirzepatide (or other weight reducing agents) in lipodystrophy. The goal of treatment may simply be to reach the healthy metabolic state which may only be possible at BMI’s that may get labeled underweight or even ‘severely malnourished’. It is important for clinicians not to use these labels when they encounter patients with lipodystrophy and work with patients to set individualized weight limits.

While our study provides preliminary data to suggest that the combined incretin pathway may lead to important metabolic benefits for patients with lipodystrophy, there are several limitations of our study. Firstly, this is only an observational cohort study where access to drug was provided clinically, and all data were collected in this setting. There was no standardized protocol for management of background medications or a data collection timetable with the noteworthy absence of quantitative dietary intake data and body composition. Secondly, the population was very heterogeneous, and patients were not selected based on predetermined criteria. Thirdly, in the absence of a randomized control group, improvements could represent Hawthorne effect or regression to the mean. Finally, it cannot be ruled out that a proportion of the observed effects may simply be a result of enhanced compliance with a once weekly administration schedule rather than the superior efficacy of GIP/GLP-1RA.

Despite these limitations, the uniformity of response across the outcomes of interest in different subgroups suggests that tirzepatide may ameliorate the stated unmet medical needs in these clinically challenging patients. There is some suggestion that tirzepatide and metreleptin (or other potential leptin agonists) may have synergistic effects. In short of restoring healthy fat tissues, such a combination may represent a therapeutic option to both replace endocrine action of fat and ameliorate consequences of ectopic spillover and/or tissue stress imposed by mismatched energy load. In conclusion, we propose that tirzepatide alone or in combination with leptin pathway agonists should be further studied in lipodystrophy with randomized controlled studies that can also help determine precise mechanisms of action.

## Data Availability

All data produced in the present study are available upon reasonable request to the authors

## Author Contributions

RM, EDF, NN, ML, RJB, and EAO cared for the patients in clinical context. EAO conceptualized the study with input from ST, RJB and MCFF. MCG, DK, ML, and RJB consented the patients for participation in each respective study. RM, MCG, DK, MCFF, ML, and RJB gathered the data. RM analyzed the data under the supervision of EAO. RM, JP, and EAO drafted the first draft of the manuscript with input from MCFF, ST and DTB . All authors reviewed and edited the final version of the manuscript and agreed to submission. EAO and RJB are the final guarantors of veracity and accuracy of data from the respective participants included from their natural history studies. EAO takes responsibility for all analyses and conduct of the study.

