## Supplemental for "Tirzepatide for Lipodystrophy"

### Supplemental Materials

| Study Name | Brief Description | Clinicaltrials.gov | IRB | Approval number |
|---|---|---|---|---|
| The LD Lync Study - Natural History Study of Lipodystrophy Syndromes | LD-Lync is a multicenter, prospective, observational natural history cohort study of genetic or acquired lipodystrophy syndromes. Patients are assessed annually and clinical data including vitals, laboratory results and anthropometric measurements are collected from patients' medical records. | NCT03087253 | UM | HUM00127427 |
| Lipodystrophy Tissue and Blood Bank study | This study is designed to collect and store tissue and blood samples from patients with lipodystrophy for research purposes. | NA | UM | HUM00062732 |
| Natural History of Disorders of Insulin Resistance | This study aimed at understanding the pathophysiology of insulin resistance and its relationship to cardiovascular disease, the molecular genetics underlying various causes of insulin resistance and diabetes mellitus, and the natural history of insulin resistance disorders, including their response to FDA approved therapies. | NCT00001987 | NIH | 76-DK-0006 |
| Observation of patients with lipodystrophy while on tirzepatide treatment | IRB exemption approval for this report | NA | UM | HUM00256460 |

**Table S1: Protocols pertaining to the participants involved in this study.** NA, Not Applicable; NIH, National Institutes of Health; IRB, Institutional Review Board; UM, University of Michigan.

| | Subtype | Diabetes Mellitus | Dyslipidemia | History of pancreatitis | MASLD or MAFLD | MASH | Cirrhosis | CAD | Hypertension | OSA | Neuropathy | Retinopathy | Acanthosis Nigricans | GERD | Gastroparesis | CKD (Stage) | PCOS | Hypothyroidism | Gout | Vitamin D3 deficiency | Osteoporosis | Iron Deficiency Anemia | Migraines | Depression | Anxiety | Chronic pain |
|---|---|---|---|---|---|---|---|---|---|---|---|---|---|---|---|---|---|---|---|---|---|---|---|---|---|---|
| #01 | FPLD1 | + | + | | + | | | | | | | | | | | I | | + | | + | | | | + | | |
| #02 | FPLD2 | + | + | | + | + | + | + | + | + | | | | | | | + | | | + | + | | | | | |
| #03 | FPLD1 | + | + | | + | | | | | | | | + | | | | + | | | | | | | + | | |
| #04 | FPLD2 | + | + | | + | | | | + | | | | + | | | | | + | | + | | + | + | | + | |
| #05 | FPLD1 | + | + | + | + | | | + | | + | + | + | + | + | | | | | | + | | + | | | | |
| #06 | FPLD3 | + | + | + | + | + | | | | | | | | | | I | + | | | | | | | | | |
| #07 | FPLD2 | + | + | | + | + | + | | | + | + | | | | + | I | + | | | + | | + | + | | + | + |
| #08 | FPLD1 | + | + | + | + | + | | | | | + | | + | + | + | | + | | | + | | | | + | | + |
| #09 | FPLD1 | + | + | | + | | | | + | | + | | + | + | | IIIa | + | | | + | | | | + | | |
| #10 | FPLD2 | + | + | | + | | | | + | | | | | + | | II | | | | | | | | | | |
| #11 | FPLD2 | + | + | | + | | | | + | | | | | + | | II | + | | + | + | + | | | + | | |
| #12 | FPLD1 | + | + | | + | | | + | + | | | | | | | IIIa | | | + | + | + | | | + | | |
| #13 | FPLD1 | + | + | + | + | | | | | + | | | | | | II | | | | | | | | | | |
| #14 | FPLD2 | + | + | | + | | | | + | + | | | + | | | I | | | | | | | + | + | + | + |
| GL1 | JDM-AGL | + | + | + | + | | | | + | | + | + | | | + | II | | | | + | | | | | | |
| GL2 | AGL | + | + | | + | + | + | | + | | | + | | | | I | | + | | | | | | | | |
| GL3 | JDM-AGL | + | + | + | + | | | | + | | | | | | | I | | | | | | | | | | |

**Table S2: Comorbidities.** *: Denotes complications that occurred during this study. AGL, Acquired Generalized Lipodystrophy; c/b, complicated by; CAD, Coronary Artery Disease; CABG, Coronary Artery Bypass Grafting; CKD, Chronic Kidney Disease; CMP, Cardiomyopathy; FPLD, Familial Partial Lipodystrophy; GERD, Gastroesophageal Reflux Disease; JDM, Juvenile Dermatomyositis; MASH, Metabolic Dysfunction-Associated Steatohepatitis; MAFLD, Metabolic Associated Fatty Liver Disease; MASLD, Metabolic Associated Steatotic Liver Disease; OSA, Obstructive Sleep Apnea; PCOS, Polycystic Ovarian Syndrome; s/p, *status-post*; tx, transplant.

| | Historical* anti-glycemic or anti-lipid medications | Pertinent medications or changed medications | | | Other, unchanged medications |
|---|---|---|---|---|---|
| | | | **Before** | **After** | |
| #01 | pioglitazone<br>metformin<br>metreleptin sc (clinical trial) | metreleptin sc- EAP<br>insulin U500 sc<br>empagliflozin<br>fenofibrate<br>rosuvastatin<br>bupropion<br>levothyroxine | 1 mg bid‡<br>80 U bid<br>25 mg qd<br>160 mg qd<br>20 mg qd<br>300 mg qd<br>300 mcg qd | d/c<br>25 U bid<br>25 mg qd<br>160 mg qd<br>20 mg qd<br>300 mg qd<br>300 mcg qd | aspirin 81 mg qd<br>ca citrate + Vit D3 qd<br>cetirizine 10 mg qd<br>estradiol patch q7d<br>medroxyprogesterone 10mg qd<br>sertraline 200 mg qd<br>vitamin D3 5000 units qd |
| #02 | pioglitazone<br>liraglutide sc<br>metreleptin sc (clinical trial) | insulin pump<br>semaglutide sc‡<br>empagliflozin<br>fenofibrate<br>metformin<br>rosuvastatin<br>spironolactone<br>bumetanide | 82 U/day<br>2.0 mg q7d<br>25 mg qd<br>145 mg qd<br>1000 mg bid<br>5 mg qd<br>100 mg qd<br>1 mg qd | 18 U/day<br>d/c<br>25 mg qd<br>145 mg qd<br>1000 mg bid<br>5 mg qd<br>25 mg qd<br>d/c | aspirin 81 mg qd<br>lactulose 30 mL bid<br>lisinopril 2.5 qd<br>magnesium 64 mg bid<br>vitamin D3 5000 units qd |
| #03 | gemfibrozil<br>liraglutide sc<br>metreleptin sc (clinical trial) | metformin<br>fenofibrate | 1000 mg bid<br>160 mg qd | 1000 mg bid<br>160 mg qd | alprazolam 0.25 mg prn |
| #04 | investigational drug or placebo sc<br>(randomized, blinded trial) | omega-3 | 1 g bid | 1 g bid | buspirone 20 mg bid<br>carvedilol 25 mg bid<br>escitalopram 20 mg qd<br>ferrous sulfate 325 mg qd<br>vitamin D3 1000 units bid |
| #05 | canagliflozin<br>dapagliflozin<br>glimepiride<br>liraglutide sc<br>pioglitazone<br>volanesorsen (clinical trial) | insulin U500 sc<br>acarbose<br>coenzyme Q10<br>empagliflozin<br>evolocumab sc<br>ezetimibe<br>fenofibrate<br>icosapent ethyl<br>metformin<br>omeprazole<br>rosuvastatin | 140 U bid<br>100 mg tid<br>150 mg bid<br>25 mg qd<br>140 mg q14d<br>10 mg qd<br>160 mg qd<br>2 g bid<br>1000 mg bid<br>20 mg bid<br>5 mg qd | 80 U bid<br>100 mg tid<br>150 mg bid<br>25 mg qd<br>140 mg q14d<br>10 mg qd<br>160 mg qd<br>2 g bid<br>1000 mg bid<br>d/c<br>5 mg qd | aspirin 81 mg qd<br>ergocalciferol 50000 units q7d<br>lisinopril 10 mg qd<br>magnesium 128 mg qd<br>ferrous sulfate 324 mg qod |
| #06 | rosiglitazone | insulin pump<br>dulaglutide sc‡¶<br>empagliflozin<br>fenofibrate<br>icosapent ethyl<br>pioglitazone | 124 U/day<br>1.5 mg q7d<br>25 mg qd<br>160 mg qd<br>2 g bid<br>15 mg qd | 111 U/day<br>d/c<br>25 mg qd<br>160 mg qd<br>2 g bid<br>15 mg qd | lisinopril 2.5 mg qd |
| #07 | exenatide sc<br>liraglutide sc<br>dulaglutide sc<br>metreleptin sc (clinical trial) | insulin U500 sc<br>metreleptin EAP sc‡<br>atorvastatin<br>empagliflozin<br>metformin XR | 125 U tid<br>10 mg qod<br>10 mg qd<br>10 mg qd<br>1000 mg bid | 60 U tid<br>d/c<br>10 mg qd<br>10 mg qd<br>1000 mg bid | biotin 1 mg qd<br>cyclobenzaprine 10 mg qd<br>duloxetine 30 mg qd<br>gabapentin 800 mg bid<br>lisinopril 5 mg qd<br>magnesium 64 mg bid<br>modafinil 200 mg qd<br>spironolactone 50 mg bid<br>tizanidine 4 mg qd<br>valacyclovir 500 mg qd<br>vitamin D3 400 units qd |

| | Historical* anti-glycemic or anti-lipid medications | Pertinent medications or changed medications | | | Other, unchanged medications |
|---|---|---|---|---|---|
| | | | **Before** | **After** | |
| #08 | metreleptin sc (clinical trial)<br>exenatide sc<br>liraglutide sc<br>semaglutide sc<br>empagliflozin<br>topiramate | insulin glargine sc<br>insulin levemir sc<br>insulin novolin R sc<br>aripiprazole<br>tube feeds g-tube | 45 U qd<br>none<br>up to 32 U ssi tid<br>2 mg qd | d/c<br>2 units qd<br>up to 10 U ssi tid<br>2 mg qd | buprenorphine patch 20mcg/h<br>cetirizine 10 mg qd<br>cromolyn 200mg qid<br>diazepam 4 mg qd<br>diphenhydramine 50 mg prn<br>duloxetine 60 mg qd<br>gabapentin 900 mg + 1200 mg<br>linaclotide 145 mcg tid<br>omeprazole 20 mg qd<br>ondansetron 4-8 mg prn<br>simethicone 80-160 mg qid prn<br>vitamin D3 5000 units qd<br>depo-testosterone 0.25mg im q7d<br>tizanidine 2 mg tid<br>tramadol 50 mg prn |
| #09 | sitagliptin<br>canagliflozin<br>dulaglutide sc | insulin U200 pump<br>empagliflozin<br>fenofibrate<br>metformin<br>omega-3 | 378 U/day<br>25 mg qd<br>145 mg qd<br>1000 mg bid<br>4 g qd | 228 U/day<br>25 mg qd<br>145 mg qd<br>1000 mg bid<br>4 g qd | amlodipine 5 mg qd<br>carvedilol 12.5 mg bid<br>lisinopril 40 mg qd<br>gabapentin 300 mg + 600 mg<br>omeprazole 20 mg qd<br>vitamin D3 1800 U qd<br>sertraline 50 mg qd |
| #10 | sitagliptin<br>dulaglutide sc<br>liraglutide sc<br>semaglutide sc, po<br>metreleptin sc (clinical trial) | metreleptin EAP sc<br>empagliflozin 25<br>metformin 1000 bid<br>omega-3<br>rosuvastatin | 10 mg qd<br>25 mg qd<br>1000 mg bid<br>2 g qd<br>5 mg qd | | aspirin 81 mg qd<br>lisinopril 10 mg qd<br>vitamin B12<br>cyclobenzaprine 10 mg qd |
| #11 | pioglitazone<br>sitagliptin<br>metreleptin sc (clinical trial) | metreleptin sc<br>ins. glargine U300 sc<br>metformin<br>miglitol<br>dapagliflozin<br>fenofibrate<br>icosapent ethyl<br>rosuvastatin | 5 mg qam<br>45 U bid<br>1000 mg bid<br>50 mg tid<br>10 mg qam<br>160 mg qd<br>1 g bid<br>10 mg qd | 5 mg qam<br>24 U bid<br>d/c<br>d/c<br>10 mg qam<br>160 mg qd<br>1 g bid<br>10 mg qd | aspirin 81 mg qd<br>calcium + vit D3 200 U bid<br>escitalopram 20 mg qd<br>magnesium 133 mg qd<br>mycophenolate mofetil 500mg bid<br>pantoprazole 40 mg qd<br>tacrolimus 0.5 mg bid<br>vitamin B12 50 mcg qd |
| #12 | liraglutide sc<br>pioglitazone<br>metformin<br>sitagliptin<br>dapagliflozin<br>ezetimibe<br>evolocumab sc<br>rosuvastatin<br>pitavastatin | semaglutide sc‡<br>alirocumab<br>icosapent ethyl<br>methylphenidate§<br>D,L-amphetamine<br>prasterone<br>colchicine<br>allopurinol | 0.5 mg q7d<br>75 mg q14d<br>1 g qd<br>none<br>5 mg bid<br>none<br>0.6 qd<br>100 mg qam | d/c<br>75 mg q14d<br>1 g qd<br>5 mg qd<br>d/c<br>50 mg qd<br>d/c<br>d/c | aspirin 81 mg qd<br>amlodipine 5 mg qd<br>febuxostat 40 mg qd<br>lorazepam 1 mg qhs<br>telmisartan 80 mg qd<br>vitamin D3 4000 units qd<br>vitamin B12 1000 mcg qd |
| #13 | none | insulin U200 pump<br>metformin<br>empagliflozin<br>rosuvastatin§<br>icosapent ethyl<br>bupropion<br>lisinopril | 150 U/day<br>1000 mg bid<br>25 mg qd<br>none<br>2 g bid<br>300 mg qd<br>5 mg qd | 33 U/day<br>d/c<br>25 mg qd<br>20 mg qhs<br>2 g bid<br>300 mg qd<br>2.5 mg qhs | vitamin D3 qd<br>vitamin B12 1000 mcg qd |

| | Historical* anti-glycemic or anti-lipid medications | Pertinent medications or changed medications | | | Other, unchanged medications |
|---|---|---|---|---|---|
| | | | **Before** | **After** | |
| #14 | metreleptin sc<br>empagliflozin<br>atorvastatin | insulin U500<br>metformin<br>canagliflozin<br>rosuvastatin<br>fenofibrate<br>omega-3<br>bupropion<br>citalopram | 270 U/day<br>2000 mg qd<br>300 mg qd<br>20 mg qd<br>145 mg qd<br>2 g bid<br>450 mg qd<br>none | d/c<br>2000 mg qd<br>300 mg qd<br>20 mg qd<br>145 mg qd<br>2 g bid<br>450 mg qd<br>15 mg qd | lisinopril 20 mg qd<br>metoprolol tartrate 50 mg bid<br>omeprazole 40 qd<br>oxycodone 15 mg tid<br>vitamin D3 1000 U qd<br>vitamin B12 2000 mcg |
| GL1 | metreleptin sc | insulin 70/30 sc<br>pioglitazone<br>atorvastatin | 50 qam + 35 qpm<br>30 mg qd<br>40 mg qd | d/c<br>30 mg qd<br>40 mg qd | losartan 100 mg qd<br>ergocalciferol 50000 IU qmonth<br>HCTZ 12.5 mg qd |
| GL2 | linagliptin<br>glimepride | metformin<br>atorvastatin<br>alogliptin<br>fenofibrate§ | 1000 mg bid<br>80 mg qd<br>25 mg<br>none | 1000 mg bid<br>80 mg qd<br>d/c<br>134 mg qd | amlodipine 10 mg qd<br>carvedilol 25 mg bid<br>furosemide 40 qd<br>losartan 100 mg qd<br>potassium chloride 20 mg qd |
| GL3 | semaglutide sc | insulin U500 sc<br>metreleptin sc<br>metformin<br>canagliflozin<br>atorvastatin<br>fenofibrate<br>ezetimibe | 1000 U/day<br>110 U bid<br>1000 mg bid<br>300 mg qd<br>40 mg qd<br>145 mg qd<br>10 mg qd | 750 U/day<br>110 U bid<br>1000 mg bid<br>300 mg qd<br>40 mg qd<br>145 mg qd<br>10 mg qd | atenolol 25 mg qd<br>calcium 1250 mg bid<br>cetirizine 5 mg qd<br>enalapril 7.5 mg bid<br>estradiol patch 2x/week<br>medroxyprogesterone loading q3months<br>omeprazole 40 mg qd<br>vitamin D3 2000 units qd |

**Table S3: Concomitant medications.** All medications are per oral (po) route unless otherwise specified. *: Last dose of medication was more than 3 months before the first dose of tirzepatide ‡: Medication has been discontinued upon initiation of tirzepatide ¶: This medication has been reinitiated upon patient's inability to obtain tirzepatide due to shortage §: This medication has been started within 3 months of tirzepatide initiation and is a potential confounder. Data beyond shortage are masked unless stated otherwise. bid, *bis in die* two times a day; d/c, discontinued; EAP, Expanded-Access Protocol[1]; HCTZ, Hydrochlorothiazide; LFT, Liver Function Tests; im, intra-muscular; IU, International Units; prn, *pro re nata* as needed; q14d, every 14 days; q7d, every 7 days; q3months, every 3 months; qam, *quaque die ante meridiem* every morning; qd, *quaque die* every day; qhs, *quaque hora somni* every night; qid, *quater in die* four times a day; qmonth, once every month; qod, *quaque altera die* every other day; qpm, every afternoon; tid, *ter in die* three times a day; sc, sub-cutaneous; ssi, sliding scale insulin; U, Units.

| | Serum Leptin (ng/mL) | | | Dexa %fat | Alanine Transaminase (U/L) | | Aspartate Transaminase (U/L) | |
|---|---|---|---|---|---|---|---|---|
| | Before metreleptin exposure | Before tirzepatide | After tirzepatide | Before | Before | After | Before | After |
| #01 | NA | NA* | 5.0 | 39.8 | 34 | 54 | 27 | 30 |
| #02 | not exposed | 8.6 | 2.6 | NA | 42 | 34 | 39 | 25 |
| #03 | not exposed | 20 | 11 | 42.0 | 45 | 25 | 63 | 14 |
| #04 | not exposed | NA | NA | 22.4 | 33 | 27 | 16 | 14 |
| #05 | not exposed | 31 | 22 | 42.6 | 24 | 61 | 17 | 49 |
| #06 | not exposed | 3.5 | 4.3 | 31.1 | 49 | 41 | 39 | 29 |
| #07 | 9.0 | NA* | 11 | 22.7 | 89 | 49 | 89 | 27 |
| #08 | not exposed | 32 | 4.3 | 35.8 | 20 | 20 | 43 | 22 |
| #09 | not exposed | 36 | NA | NA | 32 | 46 | 21 | 34 |
| #10 | 4.4 | 9.3 | NA | 14.5 | 15 | 27 | 18 | 23 |
| #11 | 16.6 | NA* | NA* | 24.7 | 28 | NA | 30 | NA |
| #12 | not exposed | 8.1 | NA | NA | 25 | 15 | 22 | 18 |
| #13 | 25 | 25 | NA | 22.0 | 28 | 14 | 19 | 12 |
| #14 | 4.6 | 23.4 | NA | 16.7 | 59 | 47 | 40 | 34 |
| GL1 | 12 | NA | NA | NA§ | 63 | 79 | 34 | 44 |
| GL2 | not exposed | 7.3 | 9.4 | NA | 48 | 35 | 34 | 30 |
| GL3 | 2.0 | NA* | NA | 14.1 | 30 | 46 | 20 | 22 |

**Table S4: Other baseline characteristics** *Patient is taking metreleptin at the time of blood draw and serum leptin level is not interpretable. §Number reported by the scanner is not interpretable due to underlying muscle disease and scar tissue. NA, Not Available

| ID | Reason for discontinuation of other GLP-1RA | Adverse effects while on tirzepatide |
|---|---|---|
| #01 | Never on GLP-1 agents | no bloating, diarrhea now and then; 1 or 2 episodes of hypoglycemia - shortly after starting 15mg - lowest 65 then 70 (doesn't remember dates) |
| #02 | Patient identified goal to come off of insulin pump, search for more potent incretin | no adverse effects |
| #03 | Never on GLP-1 agents | not many side effects except for more gas and occasional stomach sensitivity; itchy injection site |
| #04 | Never on GLP-1 agents | no adverse effects |
| #05 | Had pancreatitis on liraglutide, started on tirzepatide 3 years later | excess gas/burping, which is unpleasant, noticed slowed down GI tract - dried prunes and increased water intake |
| #06 | Alternative treatment lasting 1 month until insurance approval of more potent drug | pancreatitis due to shortage-mandated abrupt cessation , followed by Dulaglutide therapy |
| #07 | Unable to tolerate any of the GLP-1 agents, severe nausea and vomiting, and abdominal pain | upset stomach, acid reflux; 1-3 days after injection, blood sugar gets into the 50s; insulin use: from 3 times a day to 1-2 times a day, 60%, 50% or 75% less units |
| #08 | Unable to tolerate any of the agents, severe nausea, vomiting and abdominal pain | no adverse effects |
| #09 | Unable to tolerate any of the agents, severe nausea, vomiting and abdominal pain | nausea for a few days following dose switch; cannot tolerate highest dosage due to severe nausea; some episodes of hypoglycemia (50s) in middle of night; no change in insulin use |
| #10 | Could not tolerate any GLP-1 agonists given severe GERD interfering with work | severe GERD |
| #11 | Never on any GLP-1 agent | no adverse effects |
| #12 | Unable to tolerate these two agents, severe nausea, and abdominal pain | mild nausea on day #1 after injections, mild GERD |
| #13 | Never on GLP-1 agents | no adverse effects |
| #14 | Never on GLP-1 agents | no adverse events |
| GL1 | Never on GLP-1 agents | cosmetic concerns related to facial fat loss |
| GL2 | Never on GLP-1 agents | mild GERD, constipation |
| GL3 | Unable to tolerate semaglutide due to nausea | some nausea for a few days after taking the dose but is generally tolerating it well |

**Table S5: Reason for discontinuation of previous GLP-1RA and tolerance of tirzepatide.** GERD, gastroesophageal reflux disease; GLP-1RA, Glucagon-like Peptide-1 Receptor Agonists.

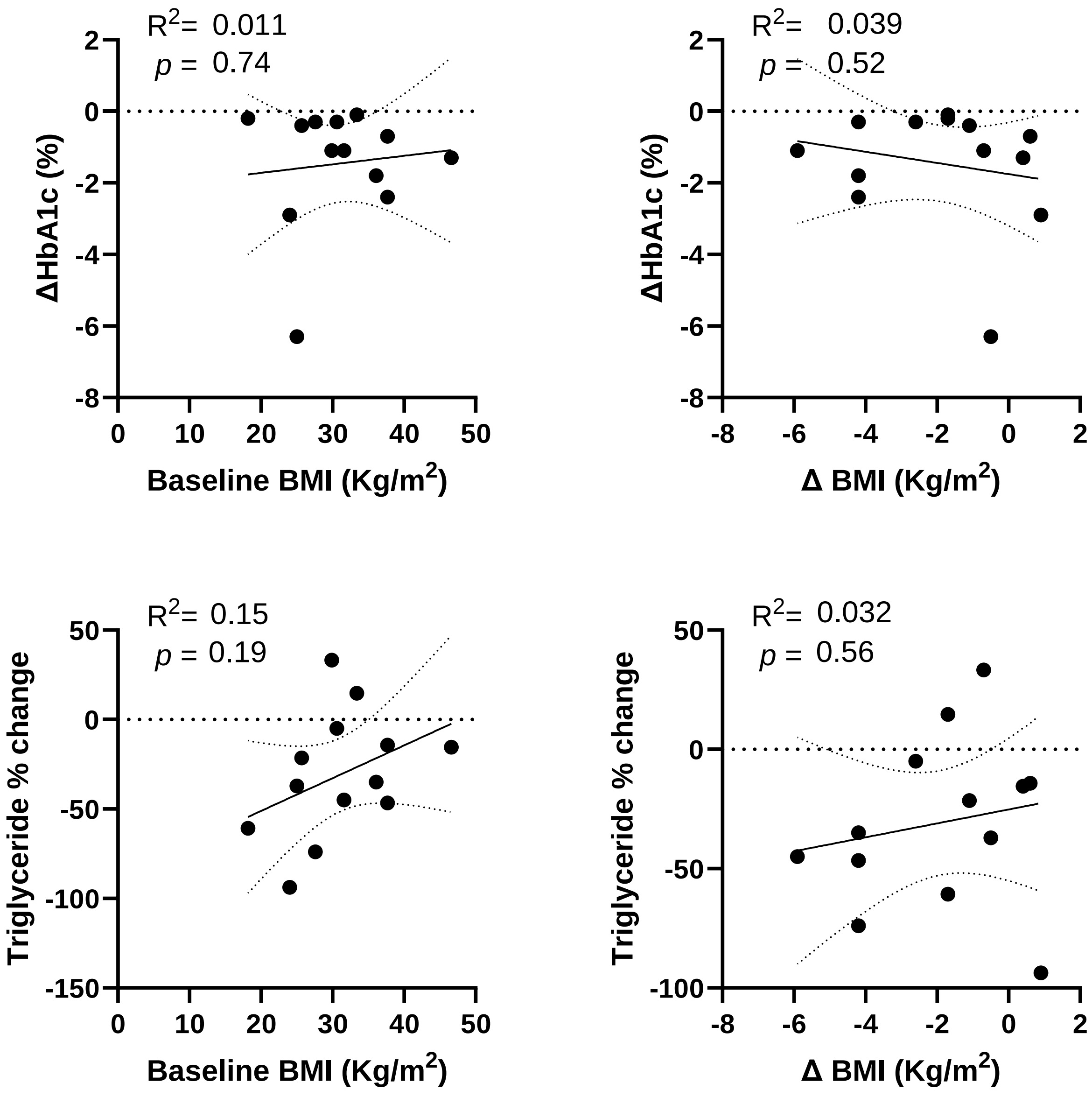


**Figure S1:** In patients with familial partial lipodystrophy, baseline BMI or change in BMI did not predict changes in HbA1c or triglycerides before and after tirzepatide.

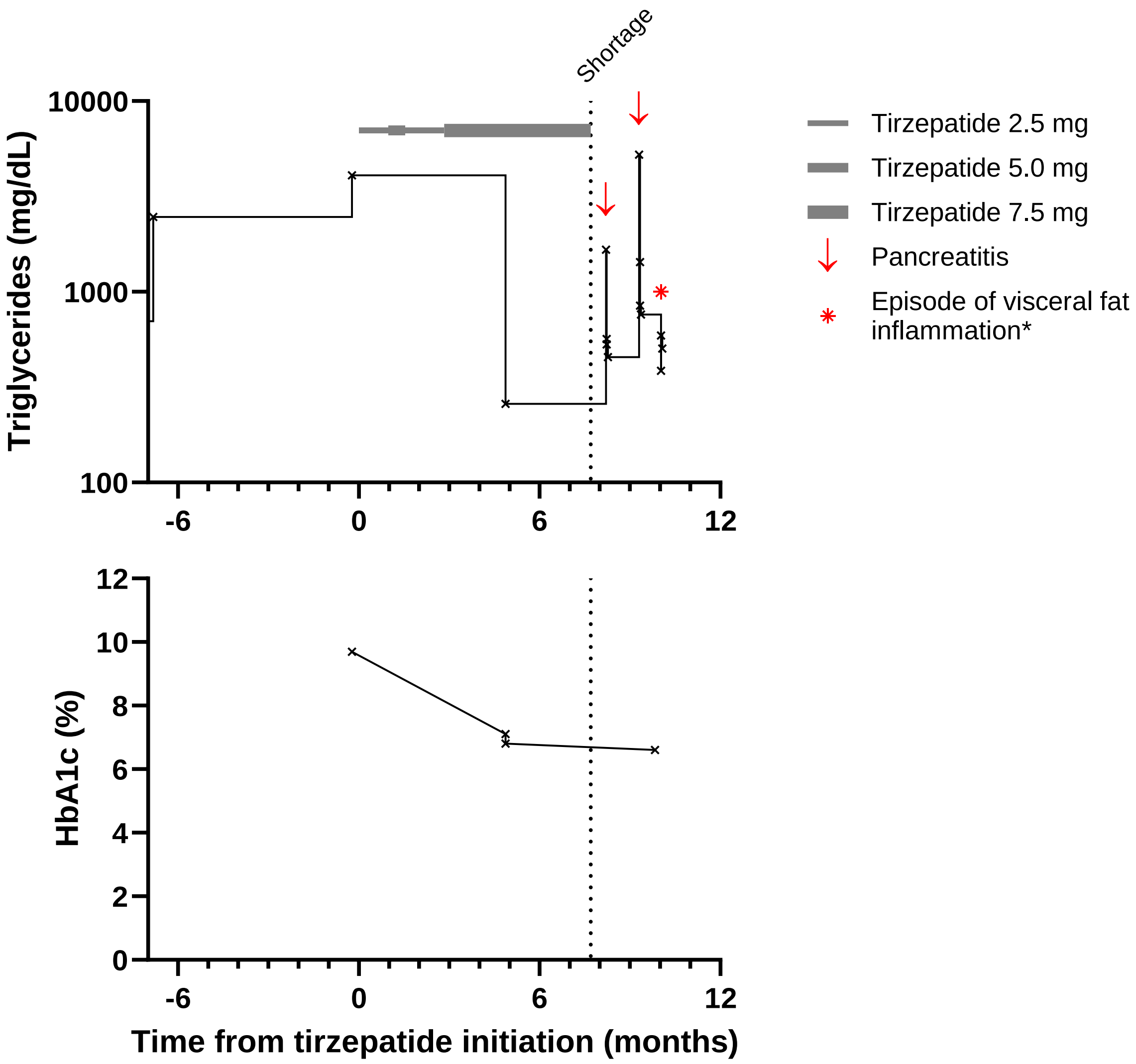


**Figure S2: Pancreatitis after drug shortage.** Patient #06 was a female with age within 35-39 years with known likely pathogenic *PPARG* missense variant (p.L178*) with a complex history including hepatosplenomegaly, neutropenia, thrombocytopenia as well as recurrent hypertriglyceridemia induced pancreatitis. Her triglycerides typically ranged from 800 to 4000 mg/dL (9-45 mmol/L) despite fenofibrate and icosapent ethyl. She received a brief course of dulaglutide 1.5 mg/week and transitioned to tirzepatide after insurance approval. After 5.5 months of tirzepatide, her triglycerides improved to an unprecedented level of 259 mg/dL (2.92 mmol/L). She was affected by a national drug shortage beginning of March 2024. She was transitioned back to dulaglutide 4.5, of which she received 1 dose, but presented to the emergency department with an episode of pancreatitis 3 weeks after being impacted by the shortage. Her triglycerides were found to have increased back up to 1660 mg/dL (18.8 mmol/L). During this admission, her dulaglutide was also discontinued citing the FDA black box warning about dulaglutide's association with pancreatitis. A month later, she had another episode of pancreatitis with triglycerides now up to 5240 mg/dL (59.2 mmol/L). *: She was admitted a third time with abdominal pain, but her lipase was normal and this episode was attributed to an acute inflammation of her visceral fat. She is still off of any incretins at the time of this writing.

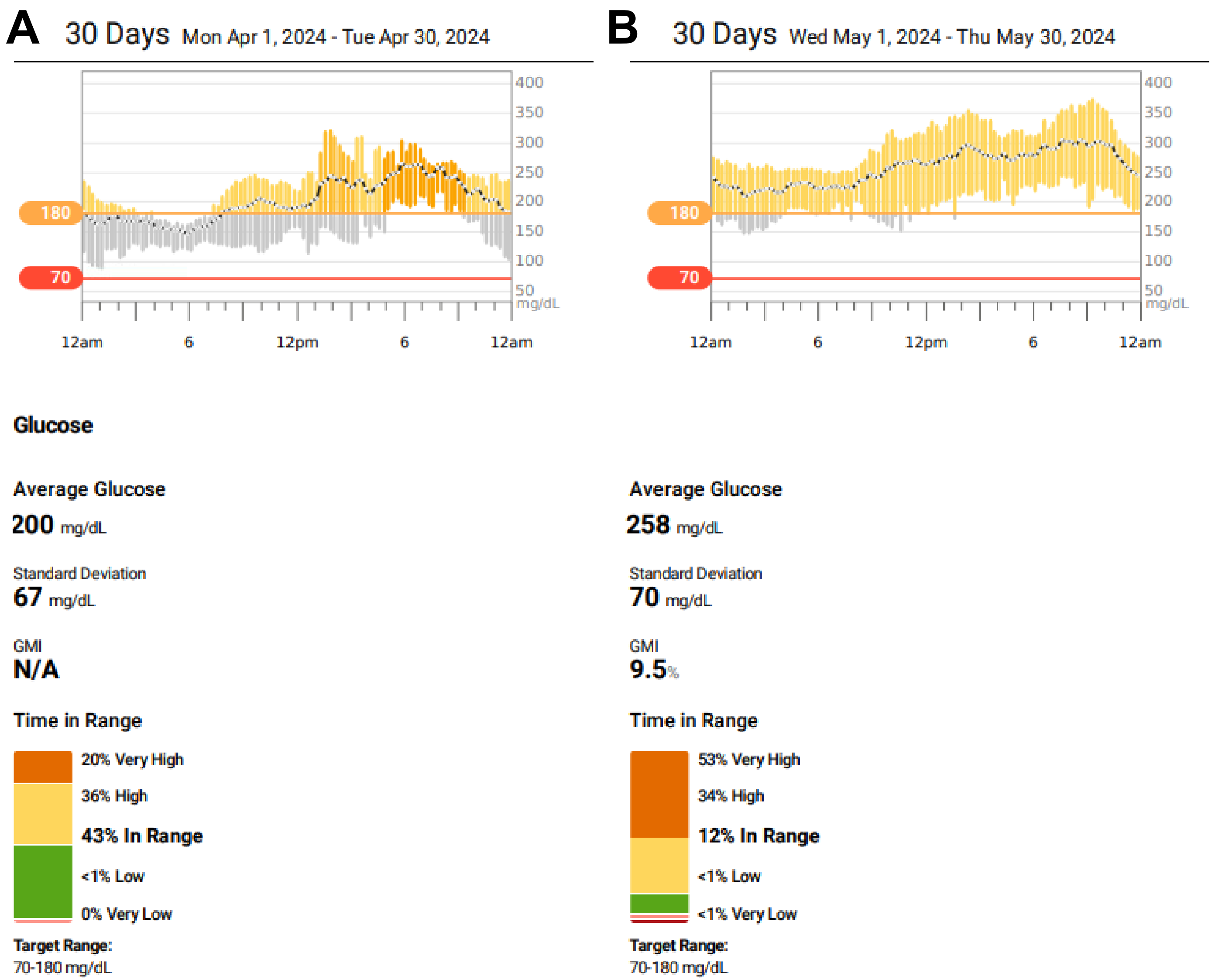


**Figure S3: Dramatic worsening of glycemic control after drug shortage.** Patient #07 was a female with age within 40-44 years range with FPLD2 (*LMNA* p.R482L) complicated by MASH induced cirrhosis who had inadequate glycemic control as evidenced by continuous glucometer (CGM) reporting time in range (TIR) of 31% and A1c 8.6%) despite insulin U500 125 units three times a day (total 375 units/day). She was on a relatively modest dose of tirzepatide (2.5 or 5.0 mg/week intermittently) as dose escalations were limited by GERD. During treatment with tirzepatide, her insulin U500 dose needed to be decreased aggressively to 60 units/day due to episodes of hypoglycemia detected by her CGM. Her CGM TIR were much improved, ranging 43%-73%. In addition, she lost 5 Kg (BMI 33.4 -> 31.5 Kg/m$^2$). Her A1c was stable at 8.5%. She was affected by drug shortage during the first week of April 2024. **Panel A** demostrates reasonably well controlled glucose during the month of April. **Panel B** demonstrates how her TIR went down to 12% in May. In addition, her triglycerides increased to 322 mg/dL (3.6 mmol/L), and end of May 2024, her insulin U500 dose needed to be readjusted back up to 375 IU/day. GMI, Glucose Management Indicator.

1. Ajluni N, Dar M, Xu J, Neidert AH, Oral EA. Efficacy and Safety of Metreleptin in Patients with Partial Lipodystrophy: Lessons from an Expanded Access Program. J Diabetes Metab 2016;7(3). DOI: 10.4172/2155-6156.1000659.
